# Novel missense variants associated with GNE myopathy

**DOI:** 10.1101/2025.07.18.25331694

**Authors:** Johanna Ranta-aho, Viviana Cetrangolo, Luca Bello, Giuliana Capece, Elena Pegoraro, Maria Francesca Di Feo, Violeta Mihaylova, Hans H. Jung, Fabien Hauw, Tanya Stojkovic, Anthony Behin, Norma Romero, Thierry Maisonobe, Matteo Lucchini, Miguel Oliveira Santos, Massimiliano Mirabella, Giorgio Tasca, Marco Savarese, Bjarne Udd, Mridul Johari

**Author notes:** Corresponding author: Mridul Johari, Full address: Harry Perkins Institute of Medical Research, 6 Verdun St, Nedlands, WA, 6009, Australia. Contributed equally to this study (Johanna Ranta-aho, Viviana Cetrangolo, Bjarne Udd, Mridul Johari).

## Abstract

GNE myopathy is a rare autosomal recessive skeletal muscle disorder characterized by progressive distal muscle weakness, typically starting in the lower legs and gradually involving proximal muscle groups. It is an autosomal recessive disease, caused by biallelic variants in *GNE*. To date, over 350 causative *GNE* variants have been reported, however, establishing genotype-phenotype correlation remains difficult due to clinical heterogeneity and limited patient numbers. In this study, we describe 20 unrelated European families with a diagnosis of GNE myopathy and biallelic *GNE* variants identified in either homozygosity or compound heterozygosity. While the majority of variants observed in our cohort have been previously reported, we identified five novel missense variants: p.(G355R), p.(A679T), p.(I709T), p.(V727G), and p.(V744I). Using *in silico* prediction tools and ACMG/AMP criteria, we classified p.(G355R) and p.(V727G) as likely pathogenic. The clinical features of our cohort were consistent with the classical presentation of GNE myopathy. Our findings contribute new genotype data and support ongoing efforts to refine variant interpretation in GNE myopathy. This study expands the mutational spectrum of *GNE* and reinforces the phenotypic consistency across diverse populations.

## 1 Introduction

Biallelic variants in the *GNE* gene, which encodes UDP-N-Acetylglucosamine 2-epimerase/N-Acetylmannosamine Kinase, cause an autosomal recessive GNE myopathy (GNEM), characterized by progressive muscle weakness, initially affecting the distal muscles of the lower limbs and gradually involving the hamstrings and other muscle groups over time [1, 2].

The *GNE* gene located on chromosome 9, consists of 13 exons and encodes for two major protein coding isoforms. The hGNE1 (NM_005476.7/ENST00000642385.2) transcript encodes for 722 amino acids (NP_005467) and is the MANE Select transcrtipt for *GNE*. The longer hGNE2 (NM_001128227.3/ENST00000396594.8), on the other hand, encodes for 753 amino acids (NP_001121699) and is designated as MANE Plus Clinical and commonly used for variant annotation [3].

To date, over 350 causative *GNE* variants have been reported [4]. These are predominantly missense variants, but insertions, deletions, nonsense, intronic, and splice site variants have also been identified. However, there are no recorded cases of GNEM patients with two null variants, and knock-out animal models have shown larval mortality, suggesting that the absence of *GNE* is lethal [5–7].

Globally, GNEM has an estimated worldwide incidence of 1 to 9 individuals per million (Orphanet; https://www.orpha.net) [4]. However, the prevalence varies across different populations due to founder effects and is likely still underestimated in some populations. Most *GNE* variants are unique to a single family, however, several founder variants in specific populations have been identified [8–12]. Most notably, these populations include the Middle Eastern (p.(M743T)), the East Asian (p.(C44S), p.(D207V), p.(V603L)), the Roma Bulgarian (p.(I618T)), and the Indian/Asian (p.(A662V), p.(V727M)) [9, 12–14].

Clinically, the typical first symptom of GNEM is the weakness of the anterior tibialis muscle, and consequent foot drop and gait disturbance [15–17]. This usually occurs in early adulthood [18]. As the disease progresses, the weakness slowly spreads to proximal muscles and upper extremities [5].

A unique feature of GNEM is the relatively spared quadriceps muscles despite the involvement of other muscles of the lower extremities [19]. The quadriceps sparing is a useful diagnostic feature, as it is hardly found in other neuromuscular disorders [20]. Typical histopathological findings from muscle biopsies of GNEM patients include fiber size variation and atrophic muscle fibers with rimmed vacuoles that relate to autophagosomal abnormality. Accumulation of phosphorylated TDP-43, p62 and beta-amyloid are also commonly found in myofibers of the patients [21]. Despite the characteristic features of GNEM, diagnosis is best established through a combination of clinical assessment, muscle imaging, and confirmatory genetic testing to identify pathogenic variants in both alleles [22–24]. Muscle biopsy is not routinely necessary anymore and is typically reserved for selected cases where the diagnosis remains unclear after initial evaluation.

While the addition of *GNE* in neuromuscular gene panels has eased the diagnosis of GNEM, establishing genotype-phenotype correlation remains challenging. This is partially due to significant variability of the disease phenotype and a relatively low number of patients [25]. The better established genotype-phenotype correlations include the predisposition to a milder phenotype associated with the variant p.(D207V) [26], and an earlier onset associated with p.(L539S) [27]. Phenotypic differences between homozygous patients and compound heterozygous patients have also been reported; patients with p.(V603L) in homozygosity tend to experience a more severe disease than patients with the same variant in compound heterozygosity [27]. The location of the variants with respect to the protein domains of *GNE* may also play a role in the observed phenotype: patients with variants in both enzymatic domains are more likely to have earlier disease onset and become non-ambulant, in comparison to patients who have both of their variants within the same domain [5, 28]. Despite these genotype-phenotype correlations, the genotype can only partially explain phenotypic variability, as significant inter- and intrafamilial variability is present in GNEM [12].

The genotypic and phenotypic diversity is also paralleled by the wide variety of proposed pathomechanisms underlying GNEM. Historically, sialic acid deficiency and consequent decreased sialylation of muscle glycoproteins has been widely accepted as the main pathomechanism associated with variants in *GNE*. However, recently, more research has emerged on the cellular processes related to *GNE*, suggesting that *GNE* expression can affect cytoskeletal organization, oxidative stress, endoplasmic reticulum stress, myogenesis, and muscle regeneration independently from hyposialylation [25].

In light of the expanding genotypic spectrum of *GNE* myopathy and growing insights into its diverse pathomechanisms, here we report 20 families from Europe where the patients are affected by GNEM and carry *GNE* variants in homozygosity or compound heterozygosity. Most of the variants in our cohort are previously reported in literature, but we also present five novel, previously unreported variants, expanding the genotypic and phenotypic spectrum of GNEM.

## 2 Methods

### 2.1 Patients and clinical examinations

We collected deep phenotyping data from 20 individuals. Diagnostic material was obtained by the referring clinicians with informed consent from the patients or their legal guardians. The study was performed according to the Declaration of Helsinki, and ethical permission was obtained through the institutional review board (HUS:195/13/03/00/11). Muscle imaging data, electrophysiological examination results (nerve conduction studies and needle electromyogram, EMG), creatine kinase (CK) measurements and cardiac function test results were obtained for most patients (Table 1). Myo-MRI and cardiac assessments were conducted in 16 patients.

**Table 1:**
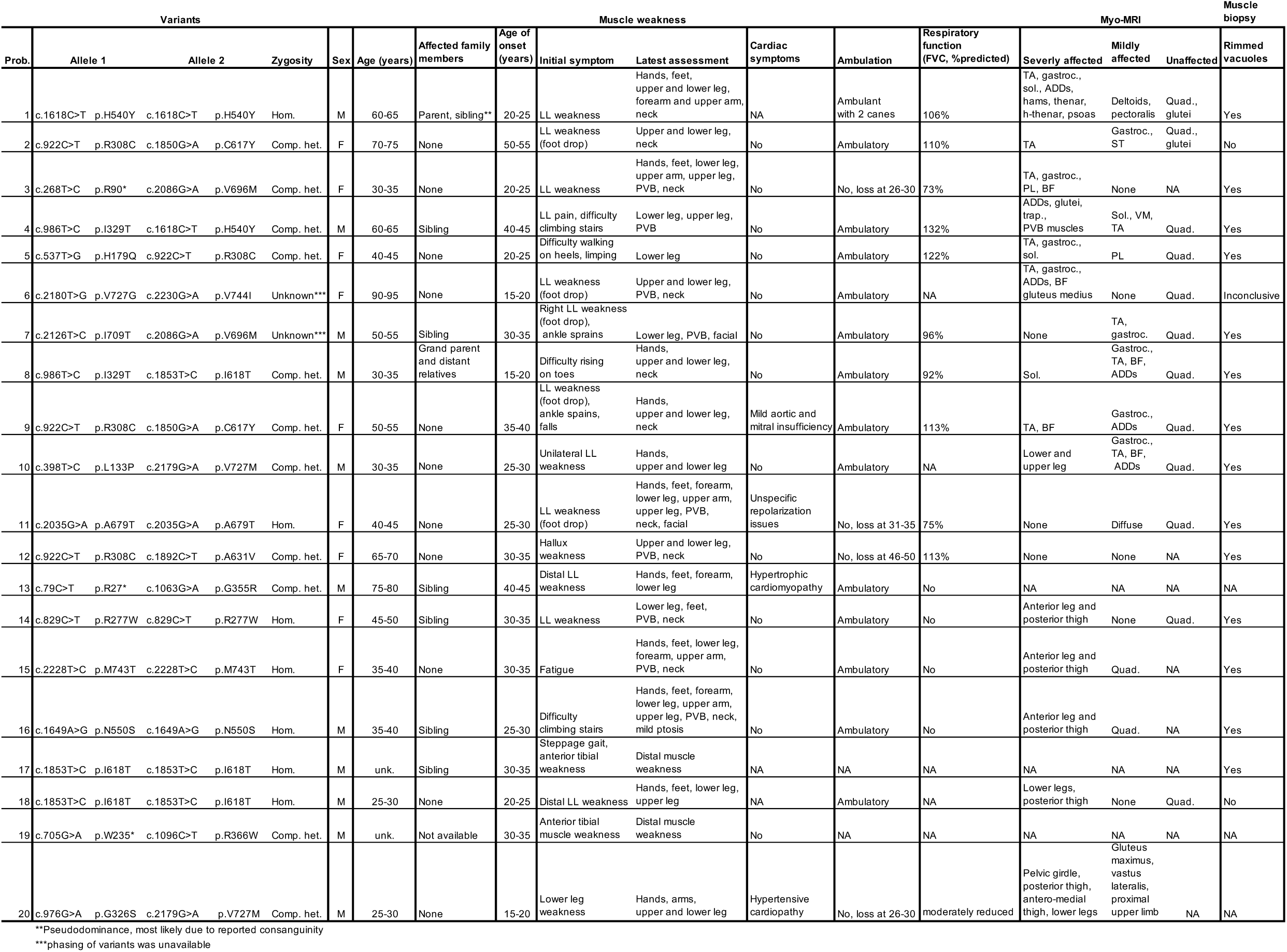
Clinical features of patient carrying biallelic *GNE* variants, included in this study. *Abbreviations:* Prob., proband; Hom., homozygous; Het., Heterozygous; M, male; F, female; unk., unknown; LL, lower limb; PVB, paravertebral; NA, not assessed; TA, tibialis anterior; gastroc., gastroncnemius; sol., soleus; ADDs, adductors; hams, hamstrings; h-thenar, hypothenar; PL, peroneus longus; BF, biceps femoris; trap, trapezius; ST, semitendinosus; VM, vastus medialis, Quad., quadriceps.

### 2.2 Molecular genetic analysis

All variants were detected by high-throughput sequencing (short read-exome or targeted gene panel) of genomic DNA obtained from the blood samples of affected individuals and confirmed by Sanger sequencing. All variants are reported using the MANE Plus Clinical transcript NM_001128227.3 (hGNE2). Pathogenicity classification of the variants was performed according to the guidelines by the American College of Medical Genetics and Genomics (ACMG) and the Association for Clinical and Genomic Science (ACGS) [29, 30]. SpliceAI was used for predicting effects of these variants on mRNA splicing. Minor allele frequencies (MAF) were obtained from the GnomAD database (version 4.1.0). AlphaMissense and AlphaFold prediction tools were also used to predict the protein structure and the variant pathogenicity [31–33].

## 3 Results

### 3.1 Clinical features

The clinical features of the 20 patients present in our cohort are summarized in Table 1. Patients 14, 15, and 16 were previously reported in an Italian cohort by Bortolani et al [34]. A positive family history was reported in 35% of the patients (7/20). All patients were followed for several years, ranging from 4-75 years, with a mean follow-up duration of approximately 20 years. The mean age at onset was 29 years, ranging from 18-50 years. The mean age at the last clinical evaluation was 50 years. Overall, most of the patients presented with muscle weakness beginning in the distal lower limbs, typically initially presenting as foot drop and weakness of the anterior tibialis. Over the course of the disease, the muscle weakness progressed to proximal lower limbs and later to the upper limbs. Within the cohort, ∼22% of the patients (4/18) had lost ambulation (Table 1), with an average time to loss of ambulation of approximately 9 years from symptom onset. Some patients also showed paravertebral (9/20), and weakness of neck muscles (11/20). Out of 17 patients with myo-MRI data, only three patients had a mildly affected quadriceps, while in 14 patients it was spared. Anterior leg and posterior thigh muscles were the most severely affected. Figure 1 shows myo-MRI of thighs and legs of patient number 20 (Table 1) with a very advanced stage of disease. Autophagic rimmed vacuoles were detected in 14/17 patients who underwent muscle biopsy. Respiratory function was evaluated in 15 patients by forced vital capacity (FVC). It was preserved or only mildly reduced in all cases. Four patients, aged between 19 and 42 years, also had unspecific cardiovascular symptoms, such as mild aortic and mitral insufficiency, hypertrophic cardiomyopathy, and hypertensive cardiomyopathy (together with type 2 diabetes, found in patient 20 at the age of 16-20).

**Figure 1:**
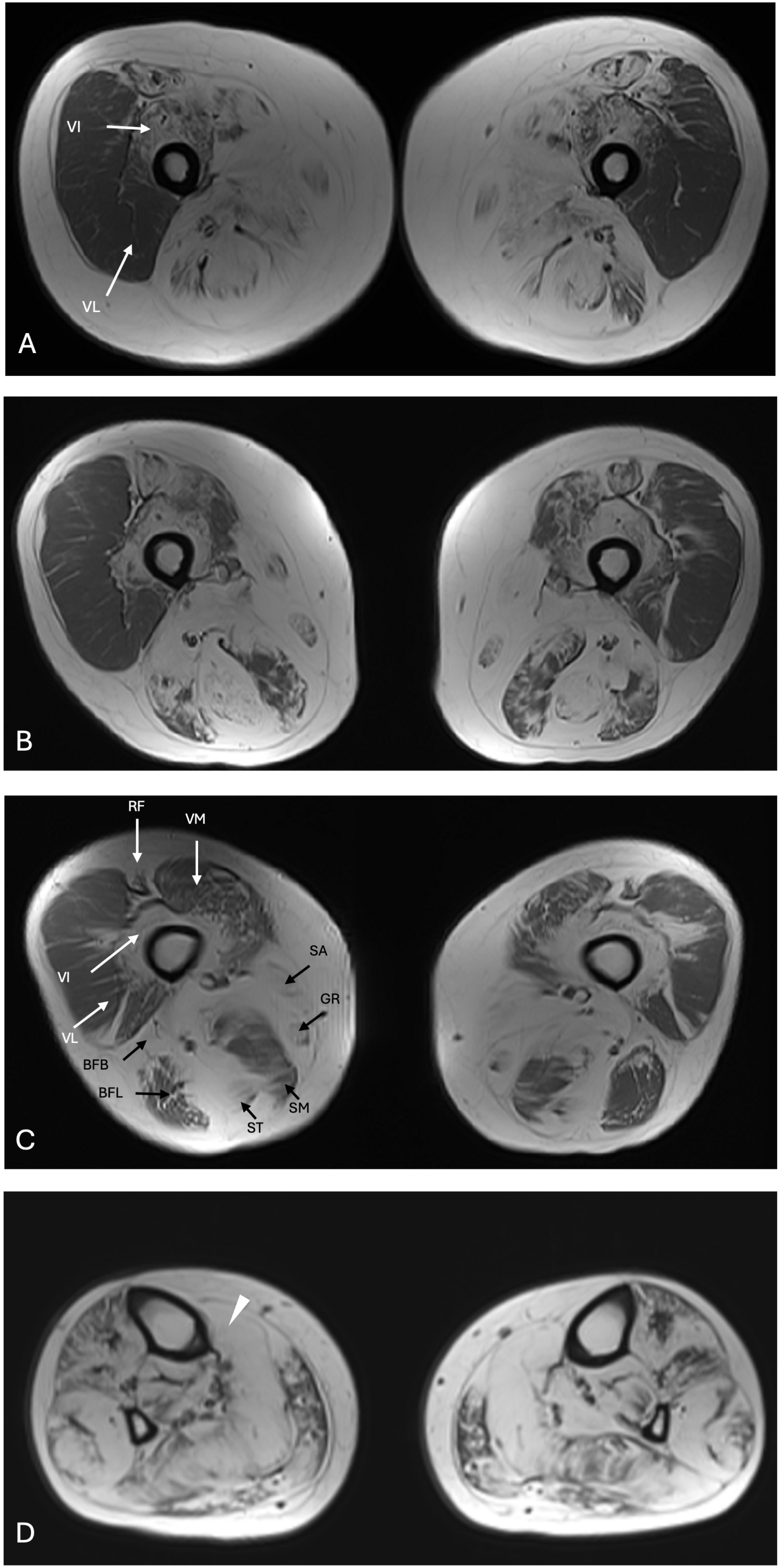
Axial T1-weighted MRI of thighs (A, B and C) and legs (D) in patient number 22 with advanced stage of disease. **A, B, and C)** The posterior and medial compartments of the thigh show severe fatty-fibrous infiltration with semitendinosus, biceps femoris brevis, sartorius, and gracilis totally replaced by fatty-fibrous tissue. The anterior compartment of the thigh is severely affected medially with rectus femoris and vastus intermedius almost totally replaced, while the vastus lateralis remains relatively spared. **D)** The fatty-fibrous infiltration of the leg muscles is diffuse and extremely severe with gastrocnemius (arrowhead) almost totally replaced. *Abbreviations:* VL, vastus lateralis; VI, vastus intermedius; VM, vastus medialis, RF, rectus femoris; SA, sartorius; GR, gracilis; ST, semitendinosus; SM, semimembranosus; BFL, biceps femoris long head; BFB, biceps femoris short head.

### 3.2 Genetics

Twenty patients from unrelated families were included in our cohort (Table 2). Fourteen patients were of European origin, one of them carrying the p.(M743T) founder variant and two of them the p.(I618T) founder variant in homozygosity. Two patients, one with parents originating from Bangladesh and the other with Comorian ancestry, carried the Indian/Asian p.(V727M) variant in compound heterozygosity with another missense variant. Altogether, in our cohort, we found 23 different *GNE* variants, including five novel, previously unreported variants. These novel variants were p.(G355R), p.(A679T), p.(I709T), p.(V727G), and p.(V744I). Seven patients carried variants in homozygosity, while 11 patients were compound heterozygous. In two patients (6 and 7), we could not further phase the variants as the patients’ parents were not available for genetic testing. The majority of the reported variants were missense, but three different heterozygous nonsense variants were also identified. The missense variants were evenly distributed across the protein structure: eight variants were located in the UDP-GlcNAc-Epimerase domain, while 12 were in the ManNAc Kinase domain. Most of the variants were rare in the general population, with a maximum frequency of less than 1 in 10,000 in the gnomAD database (version 4.1.0). The only exception were the variants p.(V727M) and p.(M743T), seen in higher frequencies (0.01241 in South Asian population, and 0.0026 in Middle Eastern population, respectively). SpliceAI scored almost all the variants lower than 0.1, and thus, they are not predicted to affect splicing. The notable exception was p.(A631V) with a SpliceAI score of 0.97.

**Table 2:**
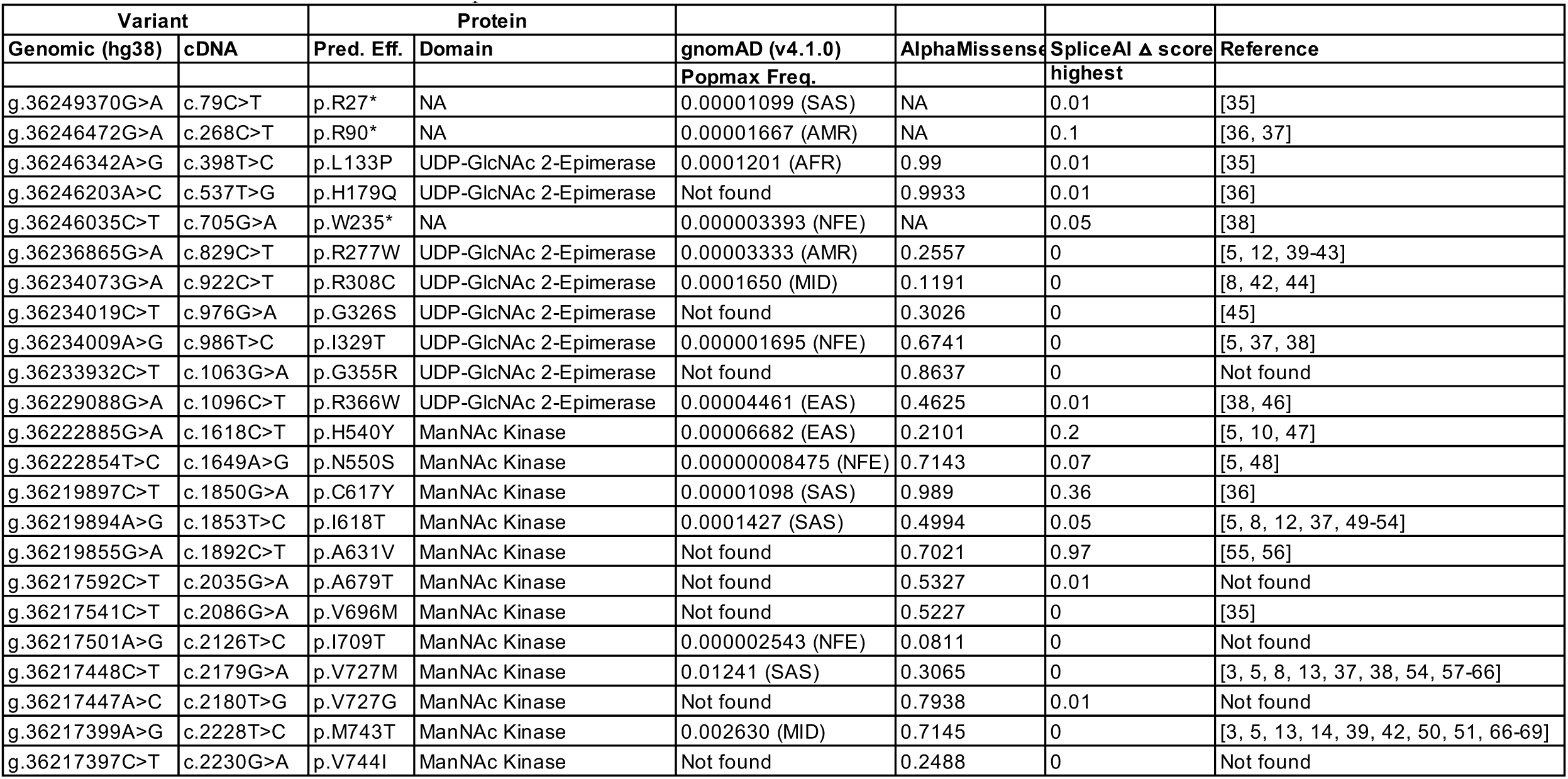
List of identified variants found in our patient cohort. *Abbreviations:* NA, not assessed; pred. eff, predicted effect; SAS, South Asian; AMR, Admixed America; AFR, African/African American; MID, Middle Eastern; NFE, non-Finnish European; EAS, East Asian.

| Variant |  | Protein |  |  |  |  |  |
| --- | --- | --- | --- | --- | --- | --- | --- |
| Genomic (hg38) | cDNA | Pred. Eff. | Domain | gnomAD (v4.1.0)<br>Popmax Freq. | AlphaMissense | SpliceAI Δ score<br>highest | Reference |
| g.36249370G>A | c.79C>T | p.R27* | NA | 0.00001099 (SAS) | NA | 0.01 | [35] |
| g.36246472G>A | c.268C>T | p.R90* | NA | 0.00001667 (AMR) | NA | 0.1 | [36, 37] |
| g.36246342A>G | c.398T>C | p.L133P | UDP-GlcNAc 2-Epimerase | 0.0001201 (AFR) | 0.99 | 0.01 | [35] |
| g.36246203A>C | c.537T>G | p.H179Q | UDP-GlcNAc 2-Epimerase | Not found | 0.9933 | 0.01 | [36] |
| g.36246035C>T | c.705G>A | p.W235* | NA | 0.000003393 (NFE) | NA | 0.05 | [38] |
| g.36236865G>A | c.829C>T | p.R277W | UDP-GlcNAc 2-Epimerase | 0.00003333 (AMR) | 0.2557 | 0 | [5, 12, 39-43] |
| g.36234073G>A | c.922C>T | p.R308C | UDP-GlcNAc 2-Epimerase | 0.0001650 (MID) | 0.1191 | 0 | [8, 42, 44] |
| g.36234019C>T | c.976G>A | p.G326S | UDP-GlcNAc 2-Epimerase | Not found | 0.3026 | 0 | [45] |
| g.36234009A>G | c.986T>C | p.I329T | UDP-GlcNAc 2-Epimerase | 0.000001695 (NFE) | 0.6741 | 0 | [5, 37, 38] |
| g.36233932C>T | c.1063G>A | p.G355R | UDP-GlcNAc 2-Epimerase | Not found | 0.8637 | 0 | Not found |
| g.36229088G>A | c.1096C>T | p.R366W | UDP-GlcNAc 2-Epimerase | 0.00004461 (EAS) | 0.4625 | 0.01 | [38, 46] |
| g.36222885G>A | c.1618C>T | p.H540Y | ManNAc Kinase | 0.00006682 (EAS) | 0.2101 | 0.2 | [5, 10, 47] |
| g.36222854T>C | c.1649A>G | p.N550S | ManNAc Kinase | 0.00000008475 (NFE) | 0.7143 | 0.07 | [5, 48] |
| g.36219897C>T | c.1850G>A | p.C617Y | ManNAc Kinase | 0.00001098 (SAS) | 0.989 | 0.36 | [36] |
| g.36219894A>G | c.1853T>C | p.I618T | ManNAc Kinase | 0.0001427 (SAS) | 0.4994 | 0.05 | [5, 8, 12, 37, 49-54] |
| g.36219855G>A | c.1892C>T | p.A631V | ManNAc Kinase | Not found | 0.7021 | 0.97 | [55, 56] |
| g.36217592C>T | c.2035G>A | p.A679T | ManNAc Kinase | Not found | 0.5327 | 0.01 | Not found |
| g.36217541C>T | c.2086G>A | p.V696M | ManNAc Kinase | Not found | 0.5227 | 0 | [35] |
| g.36217501A>G | c.2126T>C | p.I709T | ManNAc Kinase | 0.000002543 (NFE) | 0.0811 | 0 | Not found |
| g.36217448C>T | c.2179G>A | p.V727M | ManNAc Kinase | 0.01241 (SAS) | 0.3065 | 0 | [3, 5, 8, 13, 37, 38, 54, 57-66] |
| g.36217447A>C | c.2180T>G | p.V727G | ManNAc Kinase | Not found | 0.7938 | 0.01 | Not found |
| g.36217399A>G | c.2228T>C | p.M743T | ManNAc Kinase | 0.002630 (MID) | 0.7145 | 0 | [3, 5, 13, 14, 39, 42, 50, 51, 66-69] |
| g.36217397C>T | c.2230G>A | p.V744I | ManNAc Kinase | Not found | 0.2488 | 0 | Not found |

### 3.3 AlphaMissense and AlphaFold prediction

Missense variants can often lead to problems in protein folding or affect protein activity or stability. To investigate the impact of the novel missense variants identified in our study, we used AlphaMissense, a machine learning tool developed by DeepMind, widely used for predicting the pathogenicity of missense variants [31]. AlphaMissense predicted five variants as pathogenic (score of 0.57 or higher), 14 variants as uncertain significance (VUS), and four variants as benign. In accordance with the recent ClinGen PP3 criterion recommendations [70], the variants p.(G355R) and p.(V272G) were assigned supporting evidence of pathogenicity (PP3_Supporting, +1), based on their AlphaMissense scores of 0.87 and 0.80, respectively. In contrast, the variants— p.(A679T) and p.(V744I)—did not meet the threshold of PP3 criterion, with scores of 0.50 and 0.25. Additionally, the variant p.(I709T), with an AlphaMissense score of 0.08, also did not reach sufficient evidence for PP3 criterion (Table 3).

**Table 3:** Variant classification and application of American College of Medical Genetics (ACMG) guidelines for variant interpretation to previously unreported variants found in the patient cohort.

| Novel variants |  | Other allele | ACMG criteria |  |  |  |  |  |  |  |  |  | Classification |
| --- | --- | --- | --- | --- | --- | --- | --- | --- | --- | --- | --- | --- | --- |
|  |  |  | PVS1 | PP2 | PP3 | PP5 | PP4 | PM1 | PM2 | PM3 | PM5 | BP4 |  |
| c.1063G>A | p.G355R | p.R27* (Pathogenic) |  |  | Supporting |  | Supporting | Supporting | Supporting | Moderate | Supporting |  | Likely pathogenic |
| c.2035G>A | p.A679T | Homozygous |  |  |  |  | Supporting | Supporting | Supporting | Supporting | Supporting |  | VUS |
| c.2126T>C | p.I709T | p.V696M (Likely pathogenic) |  |  |  |  | Supporting | Supporting | Supporting | Moderate |  | Moderate | VUS |
| c.2180T>G | p.V727G | p.V744I (VUS) |  |  | Supporting |  | Supporting | Supporting | Supporting |  | Moderate |  | Likely pathogenic |
| c.2230G>A | p.V744I | p.V727G (Likely pathogenic) |  |  |  |  | Supporting | Supporting | Supporting | Moderate |  |  | VUS |

Furthermore, to generate and visualize the structural changes of wild-type and mutated proteins, we run AlphaFold2 [32](Figure 2). None of the novel missense variants predicted with AlphaFold2 showed a severe impact on the protein folding. However, given the structural context and the different biochemical proprieties of the mutated amino acids, these changes may still exert a pathogenic effect–possibly by impairing protein function and interactions rather than folding.

**Figure 2:**
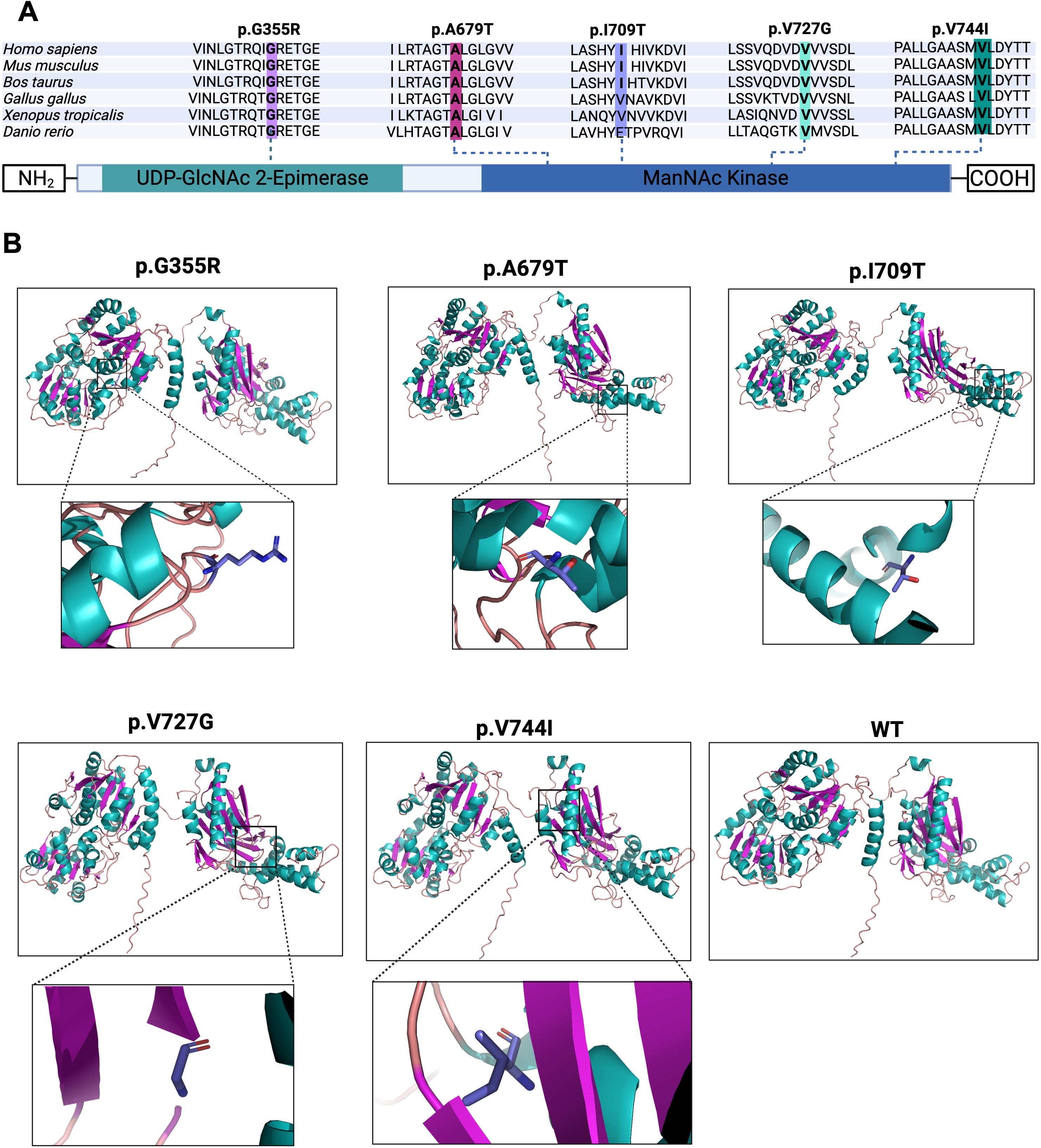
*GNE* Schematic representation and variants predicted structure. **A)** Upper: *GNE* amino acids residues and its evolutionary conserved domains, with the alignment showing comparison across several vertebrates. Middle: Schematic representation of the *GNE* structure showing the location of the variants in the UDP-GIcNAc 2-Epimerase and ManNAc Kinase domains. **B)** AlphaFold2 *GNE* novel variants predicted structure. Squares contain the details of the changed amino acid.

### 3.4 Pathogenicity classification

We classified the *GNE* variants in our cohort by predicted pathogenicity, according to the ACMG and the ACGS guidelines [29, 30]. All previously reported variants reached a classification of “pathogenic” or “likely pathogenic”, largely based on previous literature, along with additional evidence based on our cohort. As the patients in our cohort present with a phenotype compatible with GNEM, and genetically, in the majority of the patients, their *GNE* variants are observed in *trans*, we were able to add PP4_Supporting and PM3_Supporting criteria, strengthening the evidence of pathogenicity.

Similarly, we applied ACMG and ACGS guidelines to the novel *GNE* variants identified in our cohort to establish variant classification (Table 3). The variants p.(G355R) and p.(V727G) reached “likely pathogenic”, based on the observed phenotype, the low frequency of the variants, computational evidence, and location in a mutational hotspot within either the UDP-GlcNAc 2-Epimerase or the ManNac Kinase domain. The variant p.(G355R) gained additional evidence from being detected *in trans* with a pathogenic variant (PM3_Moderate +2). Finally, both variants cause a substitution of an amino acid residue where a different one has been determined to be pathogenic. Thus, PM5_Supporting was applied to p.(G355R), while PM5_Moderate was applied to p.(V727G), as there is extensive evidence supporting the pathogenicity of the variant p.(V727M) in literature [3, 5, 8, 13, 37, 38, 54, 57–66]. In contrast, the novel variants p.(A679T), p.(I709T) and p.(V744I) did not garner enough evidence to reach a classification of “pathogenic” or “likely pathogenic”, and thus, were classified as VUS.

## 4 Discussion

We report a large cohort of GNEM patients, collectively carrying 23 different variants in *GNE*. The clinical data from this cohort are highly consistent with the characteristic features of GNEM, including distal myopathy initially affecting the lower limbs, with gradual progression to proximal and upper limb muscles, relative sparing of the quadriceps, and the presence of rimmed vacuoles on muscle histopathology. Muscle imaging was typical of GNEM and provided valuable diagnostic insight. In our cohort, the proportion of patients with respiratory involvement was low, comparable to that reported in a recent Italian cohort [34]. Conversely, the proportion of patients who had lost ambulation was strikingly high, also mirroring the findings of Bortolani et al.

A few of our patients presented with comorbidities and multisystemic symptoms, including cardiovascular disease and diabetes, even at a young age. Four patients presented with a variety of cardiac symptoms, including aortic and mitral insufficiency, repolarization issues, and hypertrophic cardiomyopathy. While clear association between *GNE* variants and cardiac abnormalities has not been established [71, 72], GNEM patients present with cardiac involvement including arrythmia, valvular defects and hypertrophic cardiomyopathy at a higher frequency than the general population [72]. Interestingly, the GNEM phenotype is strikingly similar to *ACTN2*-related late-onset distal myopathy, which also begins with foot drop caused by the weakness of the tibialis muscle. However, *ACTN2* also has a long-standing association with cardiomyopathy [73]. Further, from a functional point of view, *ACTN2* and *GNE* directly interact with each other, and *GNE* variants have been shown to alter this interaction [74], which, together with the similar associated skeletal muscle phenotype, may suggest a shared disease etiology. Thus, it cannot be excluded that some of the younger patients in our cohort may develop cardiac symptoms over time. At the same time, we cannot rule out the contribution of variants in other genes to the cardiac phenotype. Similarly, studies investigating non-muscle-related comorbidities in patients with *GNE* mutations remain limited, and thus, their relation to the identified *GNE* variants is difficult to assess.

Out of the 23 *GNE* variants presented in our cohort, five were novel, previously unreported variants. Yet, three of these novel variants cause a substitution of an amino acid residue where a variant causing a different amino acid change in the same position has already been reported in literature. In particular, p.(V727M) has widely been associated GNEM, supporting the pathogenicity of p.(V727G) in our cohort [3, 5, 8, 13, 37, 38, 54, 57–66]. There are also a few reports that support the pathogenicity of variants causing an amino acid change in the same positions as p.(G355R) and p.(A679T) [43, 47, 75], however, the evidence is considerably weaker compared to p.(V727M). On the other hand, in our cohort, p.(G355R) is observed *in trans* with a null variant p.(R27*), which significantly supports its pathogenicity, and p.(A679T) is observed in homozygosity.

While recent evidence supports the pathogenicity of the novel variants p.(A679T), p.(I709T) and p.(V744I), this evidence is not robust enough to reach the formal “pathogenic” or “likely pathogenic” in ACMG classification. To further support the pathogenicity of these novel missense variants, we exploited the AlphaMissense AI tool. The scores from this tool corroborate the ACMG pathogenicity classification, except for p.(I709T), highlighting the functional impact of these variants and thus strengthening the evidence for their pathogenicity associated with GNEM phenotype of our patients. One of the AlphaMissense criteria for the pathogenicity is the structural context of a variant. Thus, depending on the location of the amino acid within the protein, the variant can affect the function of the protein. The variant p.(A679T) is classified as VUS, however, its AlphaMissense score of 0.533, its location in the functional kinase domain and in an amino acid residue where other substitutions have previously been reported as pathogenic clearly point towards pathogenicity. Conversely, the variant p.(V744I), also classified as uncertain, has a relatively low score of 0.244, decreasing its association with strong pathogenicity evidence. Finally, the considerably low score (0.08) of p.(I709T) and its subsequent classification as likely benign by AlphaMissense is probably linked to the structural location of the variant. This variant is indeed located in a structural context almost entirely classified as benign by AlphaMissense, suggesting that variants in this domain do not have a high functional impact. As we were not able to confirm that p.(I709T) existed in trans with the the pathogenic variant p.(V696M), additional evidence is required to establish its pathogenicity.

## 5 Conclusion

Overall, our results expand the genotypic and phenotypic spectrum of GNEM. Clinically, our cohort is highly consistent with previous reports of GNEM patients. A few of our patients present with comorbidities and multisystemic symptoms, including diabetes and cardiovascular disease, at a young age. Further studies, including natural history studies and patient registries will be essential to clarify the potential multisystemic impact of pathogenic *GNE* variants. Further evidence is also needed to clarify the potential impact of the three variants reported here, which were classified as VUS. This highlights how, despite significant advances in *in silico* tools for missense variant pathogenicity prediction, the sharing of clinical cases among clinicians and researchers still plays a pivotal role in gathering sufficient evidence for robust pathogenicity classification.

## Data Availability

All relevant clinical and genetic data has been shared in the manuscript. Any additional de-identified data can be made available on reasonable request, subject to appropriate data sharing agreements and ethical guidelines.

## Abbreviations

GNEM: GNE myopathy
EMG: Electromyogram
ACMG: American College of Medical Genetics and Genomics
ACGS: Association for Clinical and Genomic Science
MAF: Minor allele frequency.

## Acknowledgements

The authors would like to thank Martin Krahn for his contribution to the genetic work-up for the patients, as well as the patients and their families for cooperation in this study. Computational resources were provided by CSC – IT Center for Science, Finland.

## Author contributions

Conceptualisation of the study: MJ, BU, MS

Project administration: MJ, BU, MS

Funding acquisition: MJ, BU, MS Supervision: MJ, BU, MS

Patient samples and data collection: GC, EP, MFDF, VM, HJ, TS, AB, NR, TM, ML, MOS, MM, GT, LB, FH, and BU

Data analysis and curation: JRA, VC, MFDF, MJ

Methodology: JRA, VC, MFDF, MS, MJ

Visualisation: JRA, VC

Writing the original draft: JRA, VC

Review and editing of the manuscript: all co-authors

